# Immunogenicity of an adjuvanted, combination inactivated rabies-vectored, Lassa fever vaccine in healthy adults: interim results of a first-in-human Phase 1 trial

**DOI:** 10.1101/2025.10.16.25338178

**Authors:** Justin R Ortiz, Drishya Kurup, Adam C Kaufman, Sabrine Ben Hamed, Allison M LaRocco, Kirsten E Lyke, Sarah M Litts, Shrimati Datta, Yuanyuan Liang, Megan F McGilvray, Jeannie A Losignor, Jennifer J Oshinsky, Kari V Palmer, Ifayet PL Johnson-Mayo, Gabrielle Scher, Uthra Balakumar, Anisha Chandwani, Rekha R Rapaka, Meagan E Deming, Joel V Chua, Christoph Wirblich, Wilbur H Chen, Kathleen M Neuzil, Marcela F Pasetti, Matthias J Schnell

## Abstract

We conducted a randomized, controlled Phase 1 trial of an inactivated rabies virus-vectored Lassa virus glycoprotein complex (GPC) vaccine (LASSARAB) formulated with a synthetic oil-in-water TLR-4 agonist adjuvant (3D-6acyl PHAD-SE). In dose-escalating cohorts, participants received two intramuscular doses of LASSARAB containing 700 (n=15), 1400 (n=15), or 2800 (n=14) relative units of antigen, as quantified by ELISA using recombinant GPC as a standard, or a licensed rabies vaccine control (n=10), administered 28 days apart. This interim analysis reports the primary objective of safety evaluation and the secondary objective of immunogenicity assessment through Day 61. All doses were well tolerated. After two doses, Lassa GPC ELISA seroconversion rates were 100% in all LASSARAB groups and 0% in the control group. Rabies glycoprotein ELISA seroconversion and neutralizing antibody seroprotection were both 100% across all groups, including controls. This first-in-human study demonstrates that LASSARAB+3D-6acyl PHAD-SE is a well-tolerated, highly immunogenic, combination vaccine, supporting further clinical evaluation. ClinicalTrials.gov identifier: NCT06546709.

---

Lassa fever is a zoonotic disease endemic to West Africa caused by the Lassa virus.^1^ The World Health Organization (WHO) estimates that there are up to 300,000 infections and 5,000 deaths annually,^2^ although the true burden is likely higher because of under-diagnosis and limited surveillance.^3,4^ Transmission to humans occurs primarily through contact with food or household items contaminated by rodent urine or feces, although person-to-person spread can also occur via contaminated objects.^1^ Approximately 80% of infections are asymptomatic;^5^ however, when symptomatic, clinical manifestations range from mild febrile illness to hemorrhagic fever and death.^3,4^ Sensorineural hearing loss develops in about 25% of survivors and may result in permanent deafness.^6,7^ When infection occurs late in pregnancy, more than 80% of cases result in maternal death or fetal loss.^1,8^ The overall case-fatality rate is approximately 1%, but increases to about 15% among hospitalized patients with severe disease.^1^ No licensed vaccines are currently available.

The immune mechanisms that protect against severe Lassa fever are not fully understood. Convalescent serum from survivors has not proven effective in clinical studies,^9–11^ but monoclonal antibodies derived from survivors’ B cells have shown promise in animal models.^10,11^ Survivors typically mount an early, high-titer IgG response to the virus glycoprotein complex (GPC), which mediates cellular entry. Preclinical studies suggest that antibody-mediated protection may depend more on binding and Fc receptor-mediated effector functions than on classical neutralizing activity.^12–14^ In parallel, survivors generate robust, polyfunctional T-cell responses, particularly CD8⁺ T cells producing interferon-γ and TNF-α against GPC and the viral nucleoprotein, while maintaining relatively low systemic inflammation.^12,15^ Early seroconversion and control of viremia are associated with survival.^16,17^

Because Lassa fever has epidemic potential and poses major challenges for under-resourced health systems, global health agencies have prioritized Lassa vaccine development.^18^ The World Health Organization (WHO) lists the virus as a top-priority pathogen in its *R&D Blueprint for Action to Prevent Epidemics*, while the Coalition for Epidemic Preparedness Innovations (CEPI) has emphasized the urgent need for a safe, effective, and programmatically suitable vaccine for endemic regions.^2,19^ In parallel, WHO has underscored the value of combination vaccine approaches,^20,21^ recognizing their potential to protect against multiple pathogens within a single product and to simplify vaccine delivery in resource-limited settings. Building on these priorities, scientists at Thomas Jefferson University developed an inactivated, attenuated rabies-vectored combination vaccine expressing both the rabies glycoprotein and the Lassa virus GPC (LASSARAB).^20,21^ Adjuvanted LASSARAB showed protective efficacy in preclinical studies, preventing lethal Lassa virus challenge in animal models.^20,21^ An additional advantage of this platform is its inclusion of rabies glycoprotein, which could provide immunity to rabies as part of a combination vaccine.

In this study, we conducted a Phase 1, first-in-human, randomized, controlled trial of LASSARAB formulated with 3D-6acyl PHAD-SE (aPHAD-SE), a toll-like receptor-4–activating oil-in-water emulsion adjuvant. The primary objective was to assess the safety and reactogenicity of escalating doses of LASSARAB+aPHAD-SE. Secondary objectives were to evaluate the antibody responses to both Lassa and rabies virus antigens.

## RESULTS

### Clinical trial design and participants

There were four vaccine groups (**Table 1**). Two groups received a single injection of LASSARAB+aPHAD-SE. Group A received 700 rU of antigen with 5 µg of adjuvant, and Group B received 1400 rU of antigen with 5 µg of adjuvant. Group C received a high dose via two concurrent injections of the medium-dose preparation (2800 rU of antigen and 10 µg of adjuvant in total). Group D, the control group, received the licensed inactivated rabies vaccine (Imovax Rabies, Sanofi Pasteur, Lyon, France) containing at least 2.5 international units (IU) of rabies virus antigen per 1.0 mL dose. To maintain blinding for Group C, a subset of control recipients also received a concurrent injection of normal saline. Vaccines were administered intramuscularly on Days 1 and 29, following a dose-escalation protocol in which a sentinel subset of each dose group and the control group was vaccinated and monitored for safety before the remaining participants in that group and the next sentinel cohort were enrolled **(Supplemental Table 1)**. For this protocol-defined interim analysis, safety and immunogenicity were monitored through Day 61.

**TABLE 1.** TREATMENT GROUPS AND DESCRIPTION OF PARTICIPANTS.

|  | **Group A**<br>**(n=15)** | **Group B**<br>**(n=15)** | **Group C**<br>**(n=14)** | **Group D**<br>**(n=10)** | **Overall**<br>**(n=54)** |
| --- | --- | --- | --- | --- | --- |
| **Description** | **Low Dose** | **Medium Dose** | **High Dose** | **Control** |  |
| Age, mean (standard deviation) | 34.0 (8.9) | 32.3 (6.5) | 31.6 (7.3) | 31.8 (6.4) | 32.5 (7.3) |
| BMI, mean (standard deviation) | 26.7 (3.9) | 25.5 (3.5) | 24.1 (4.1) | 24.4 (4.0) | 25.3 (3.9) |
| Sex, n (%) |  |  |  |  |  |
| Female | 8 (53.3%) | 6 (40.0%) | 5 (35.7%) | 5 (50.0%) | 24 (44.4%) |
| Male | 7 (46.7%) | 9 (60.0%) | 9 (64.3%) | 5 (50.0%) | 30 (55.6%) |
| Ethnicity, n (%) |  |  |  |  |  |
| Hispanic | 2 (13.3%) | 2 (13.3%) | 0 (0.0%) | 2 (20.0%) | 6 (11.1%) |
| non-Hispanic | 13 (86.7%) | 13 (86.7%) | 14 (100.0%) | 8 (80.0%) | 48 (88.9%) |
| Race, n (%) |  |  |  |  |  |
| American Indian or Alaskan Native | 1 (6.7%) | 0 (0.0%) | 0 (0.0%) | 2 (20.0%) | 3 (5.6%) |
| Asian | 1 (6.7%) | 1 (6.7%) | 3 (21.4%) | 3 (30.0%) | 8 (14.8%) |
| Native Hawaiian or other Pacific Islander | 0 (0.0%) | 0 (0.0%) | 0 (0.0%) | 0 (0.0%) | 0 (0.0%) |
| Black or African American | 2 (13.3%) | 2 (13.3%) | 1 (7.1%) | 2 (20.0%) | 7 (13.0%) |
| White | 11 (73.3%) | 12 (80.0%) | 9 (64.3%) | 6 (60.0%) | 38 (70.4%) |
| Other | 0 (0.0%) | 0 (0.0%) | 0 (0.0%) | 0 (0.0%) | 0 (0.0%) |
| Unknown | 0 (0.0%) | 0 (0.0%) | 0 (0.0%) | 1 (10.0%) | 1 (1.9%) |

**Note:** Three participants reported two or more races.

The interim trial period extended from January 22, 2025, through June 24, 2025, when all participants completed Day 61. Fifty-four healthy adults aged 18 through 50 years were enrolled and randomized: 15 participants in Group A (low dose), 15 in Group B (medium dose), 14 in Group C (high dose), and 10 in Group D (control) **(Figure 1)**. Completion of the second vaccination occurred in all participants in Groups A, B, and C, and in 9 of 10 (90%) participants in Group D. All enrolled participants completed safety and immunogenicity monitoring through Day 61. One Group D participant opted out of the second vaccination for reasons unrelated to the vaccine but remained under monitoring.

**FIGURE 1.**
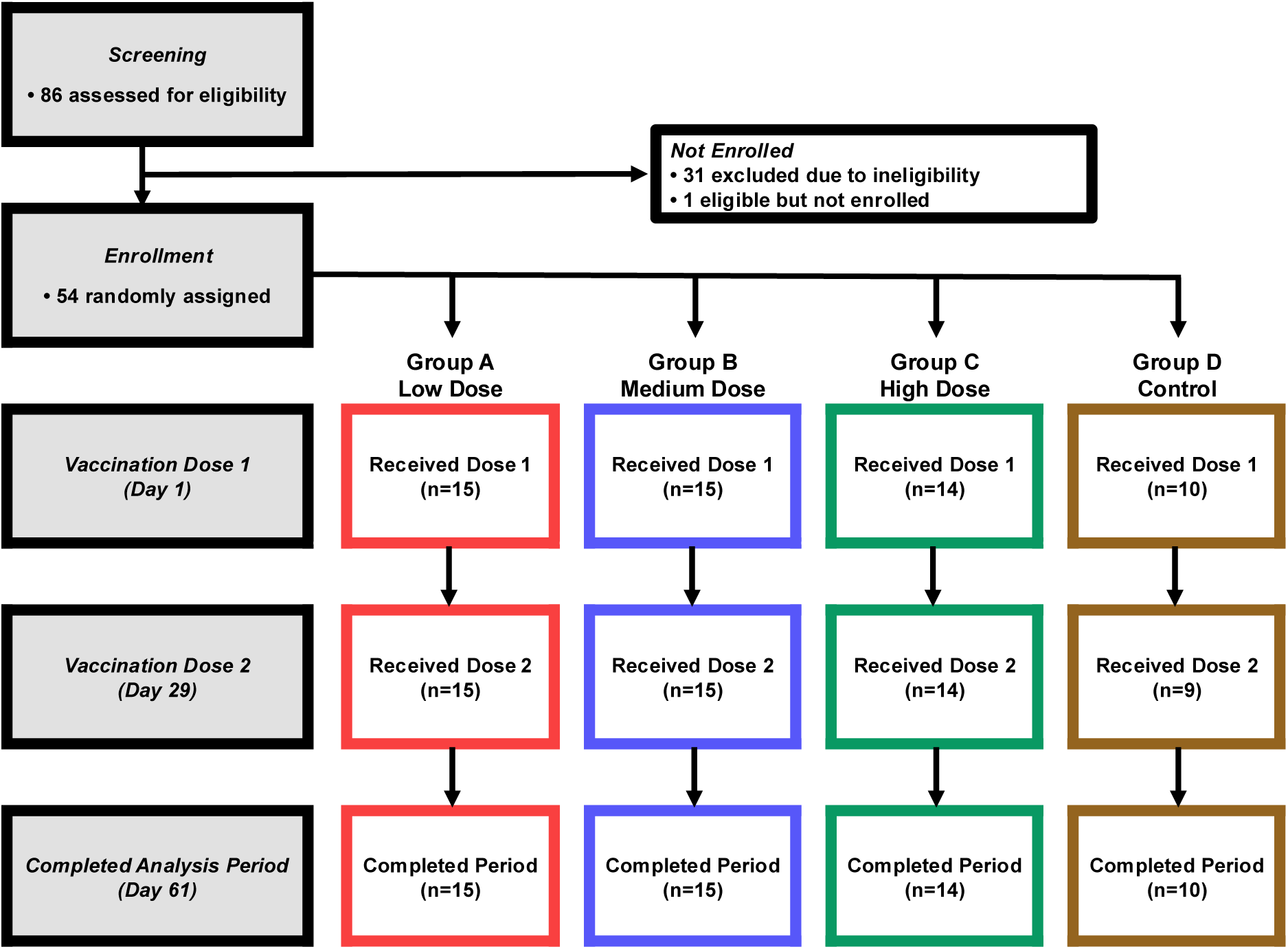
CONSORT DIAGRAM **Note:** Reasons for exclusion due to ineligibility included failed hearing test (n=13), inability to comply with study procedures (n=11), exclusionary medication (n=2), unacceptable screening laboratory results (n=2), planned allergy treatment (n=1), unacceptable screening vital signs (n=1), prior rabies vaccination (n=1), acute illness (n=1), and failed rabies screening test (n=1). Two participants had more than one reason for ineligibility. One Group D participant declined the second vaccination for reasons unrelated to the vaccine but remained under monitoring through Day 61. **Color key**: red = Group A (low dose), blue = Group B (medium dose), green = Group C (high dose), and brown = Group D (control).

Overall, 44.4% of participants were women, 13.0% were Black or African American, and 11.1% were Hispanic. The mean age was 32.5 years, and the mean body mass index was 25.3. Demographic information is presented in **Table 1**.

### Lassa virus GPC antibody responses

Baseline Lassa virus-GPC serum IgG ELISA geometric mean titers (GMTs) were low (≤2.1 IU/mL), and no significant response was detected on Day 8 in any group (**Figure 2 and Supplemental Table 2**). By Day 29, significant increases in GMTs from baseline were observed in the LASSARAB-containing vaccine groups: 8.8 IU/mL (95% CI: 3.3–23.6) for Group A, 6.4 IU/mL (95% CI: 4.0–10.3) for Group B, and 13.4 IU/mL (95% CI: 9.6–18.9) for Group C, but not in the control Group D (1.4 IU/mL; 95% CI: 0.7–2.7). On Day 29, Lassa virus-GPC antibody seroconversion (≥4-fold rise from baseline) was observed in the LASSARAB-containing vaccine groups: 60.0% (95% CI: 32.3–83.7) for Group A, 33.3% (95% CI: 11.8–61.6) for Group B, and 64.3% (95% CI: 35.1–87.2) for Group C, but not in the control Group D (0.0%; 95% CI: 0.0–33.6).

**FIGURE 2.**
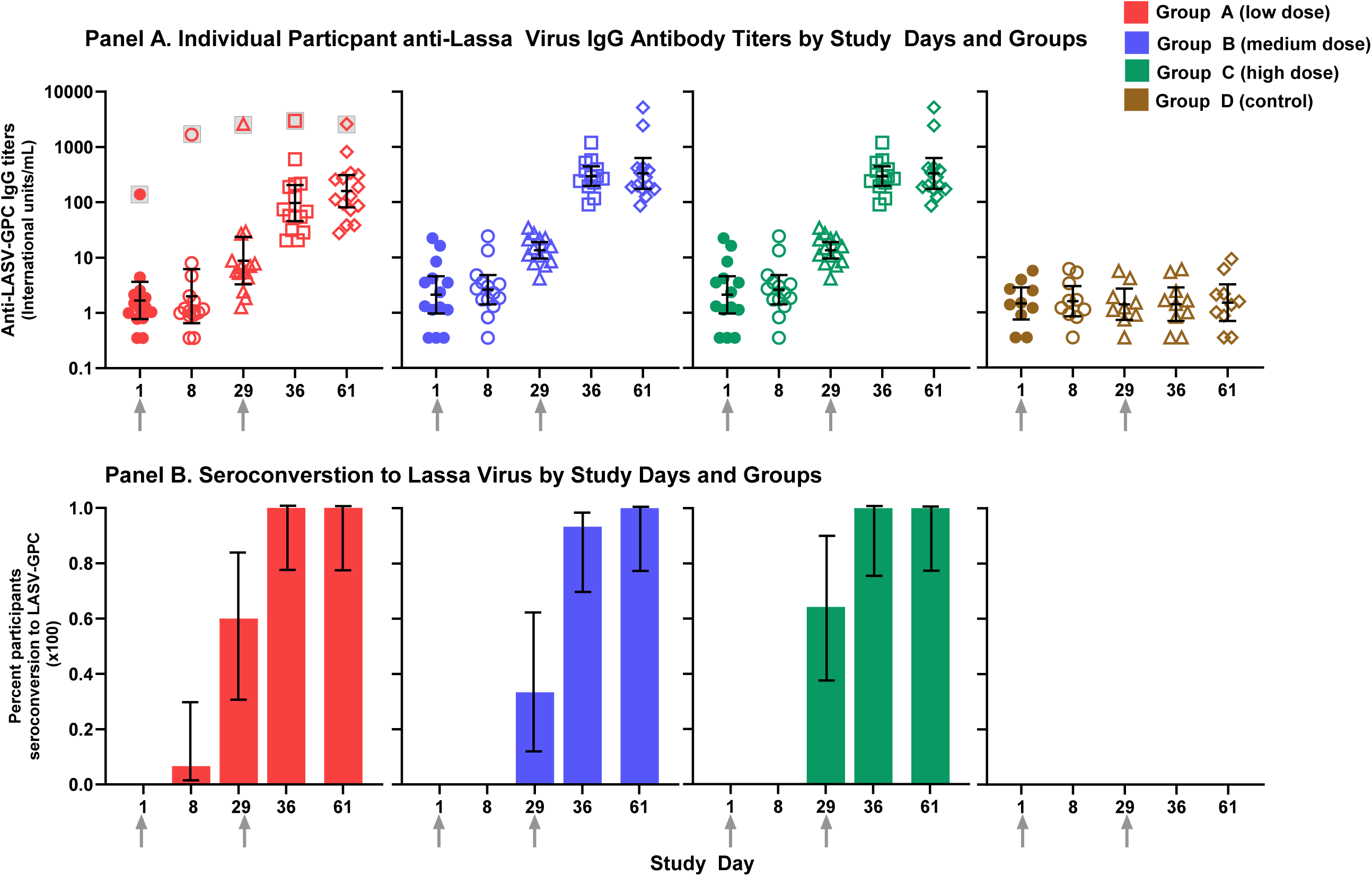
LASSA VIRUS GLYCOPROTEIN COMPLEX ANTIBODY RESPONSES BY ENZYME-LINKED IMMUNOSORBENT ASSAY (ELISA) ACROSS STUDY GROUPS AND DAYS **Note:** **Panel A.** Lassa virus glycoprotein complex (GPC) antibody titers measured by enzyme-linked immunosorbent assay (ELISA), shown by study arm and day. Individual symbols represent participant values; bars indicate geometric mean titers with 95 % confidence intervals (CIs). Grey arrows indicate Dose 1 at Day 1 and Dose 2 at Day 29; immune assay blood draws preceded vaccination on each day. Symbols denote study day: solid circles = Day 0, open circles = Day 8, triangles = Day 29, squares = Day 36, and diamonds=Day 61. One Group A participant had an elevated baseline Lassa antibody titer (138.2 IU/mL) and maintained the highest titer in that group at every time point (grey box around symbol). **Panel B.** Proportion of participants in each study arm with Lassa virus GPC seroconversion, defined as a ≥4-fold rise from Day 1 titer, by study day. Seroconversion is not applicable to Day 1. Bars show mean values with 95% confidence intervals. Grey arrows indicate Dose 1 at Day 1 and Dose 2 at Day 29; immune assay blood draws preceded vaccination on each day. **Color key:** red = Group A (low dose), blue = Group B (medium dose), green = Group C (high dose), and brown = Group D (control).

After the second vaccination, GMTs remained elevated through Day 61 in the LASSARAB-containing vaccine groups: 158.8 IU/mL (95% CI: 81.1–310.9) for Group A, 135.4 IU/mL (95% CI: 87.6–209.3) for Group B, and 331.0 IU/mL (95% CI: 174.0–629.6) for Group C, while remaining at baseline levels in the control Group D (1.5 IU/mL; 95% CI: 0.7–3.2). All LASSARAB-containing vaccine groups achieved seroconversion by Day 61.

There was no evidence of a dose response among LASSARAB-containing vaccines at any time point, though one participant in Group A had a high baseline Lassa virus antibody titer (138.2 IU/mL) suggesting prior exposure. To assess whether this influenced outcomes, we conducted a post hoc analysis excluding this participant. After exclusion, baseline Lassa virus IgG GMT in Group A was 1.2 IU/mL (95% CI: 0.8–1.8), with subsequent GMTs of 1.2 IU/mL (95% CI: 0.8–2.0) on Day 8, 5.9 IU/mL (95% CI: 3.5–9.7) on Day 29, and 130.0 IU/mL (95% CI: 74.3–227.4) on Day 61. Comparing these post hoc results to Groups B and C at Day 29 to evaluate the potential best dose for a single-dose vaccine, we found Group A responses were statistically lower than those of Group C, while no statistical difference was observed between Groups B and C. At Day 61, analysis of the two-dose regimen showed statistically comparable immune responses across all LASSARAB-containing vaccine groups.

### Rabies virus glycoprotein antibody responses

Baseline rabies virus glycoprotein serum IgG ELISA GMTs were low (≤0.1 IU/mL), and only minimal, non-significant increases were detected on Day 8 in all four groups (**Figure 3 and Supplemental Table 3**). By Day 29, GMTs increased significantly from baseline in all groups: 5.9 IU/mL (95% CI: 3.5–9.8) for Group A, 8.6 IU/mL (95% CI: 5.8–12.8) for Group B, 19.0 IU/mL (95% CI: 12.6–28.7) for Group C, and 1.7 IU/mL (95% CI: 0.9–3.3) for the control Group D. After the second vaccination, GMTs increased further, peaking at Day 36: 45.0 IU/mL (95% CI: 29.5–68.6) for Group A, 73.2 IU/mL (95% CI: 45.6–117.5) for Group B, 170.3 IU/mL (95% CI: 128.3–226.2) for Group C, and 26.5 IU/mL (95% CI: 11.8–59.5) for the control Group D. Day 61 GMTs had declined slightly. On Day 29, seroconversion (defined as a ≥4-fold rise) was observed in 100% of participants across all groups, and this rate remained constant at 100% through Day 61.

**FIGURE 3.**
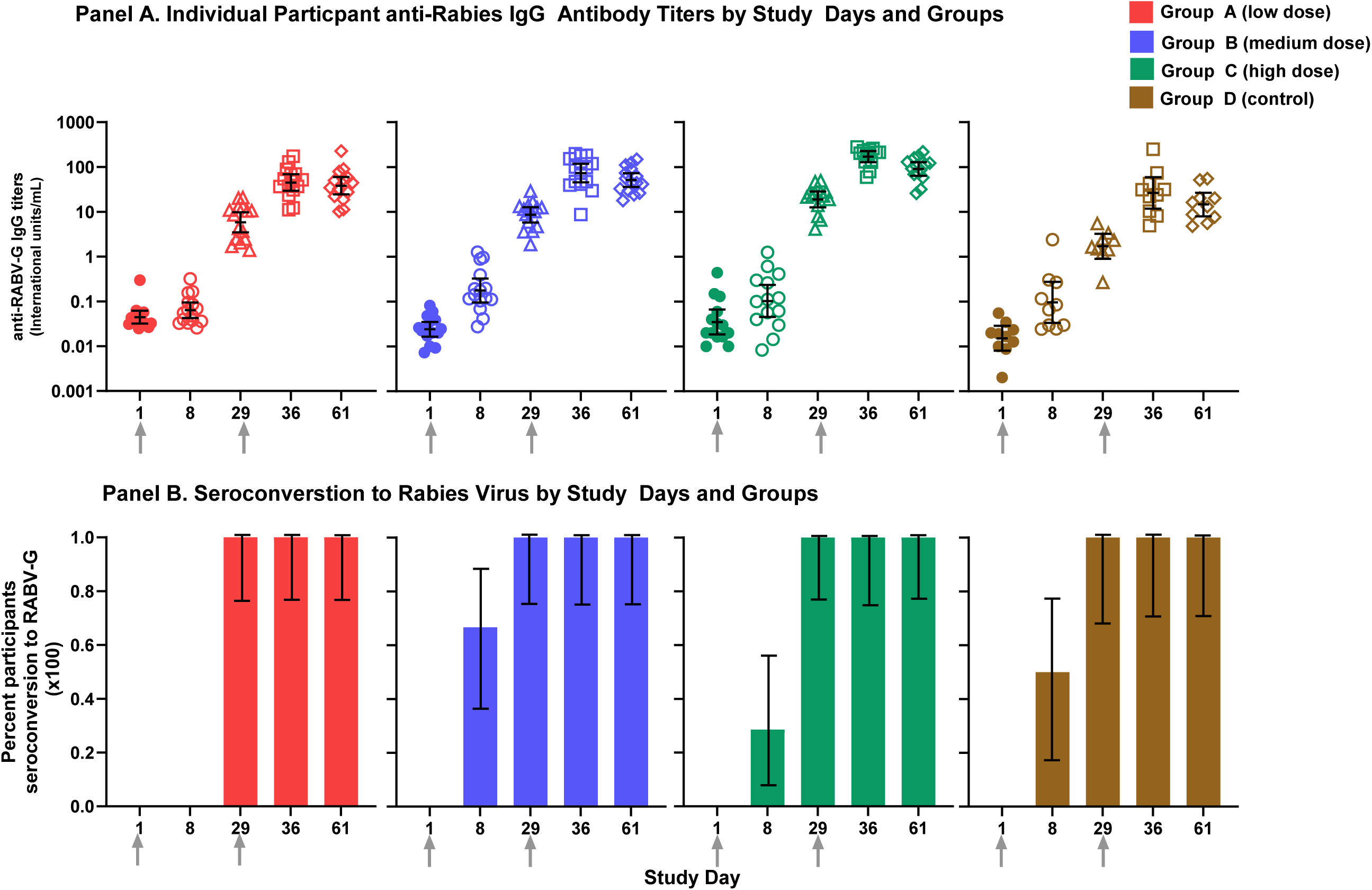
RABIES VIRUS GLYCOPROTEIN ANTIBODY RESPONSES BY ENZYME-LINKED IMMUNOSORBENT ASSAY (ELISA) ACROSS STUDY GROUPS AND DAYS **Note:** **Panel A.** Rabies virus glycoprotein antibody titers measured by enzyme-linked immunosorbent assay (ELISA), shown by study arm and day. Individual symbols represent participant values; bars indicate geometric mean titers with 95 % confidence intervals (CIs). Grey arrows indicate Dose 1 at Day 1 and Dose 2 at Day 29; immune assay blood draws preceded vaccination on each day. Symbols denote study day: solid circles = Day 0, open circles = Day 8, triangles = Day 29, squares = Day 36, and diamonds=Day 61. **Panel B.** Proportion of participants in each study arm with seroconversion, defined as a ≥4-fold rise from baseline titer, by study day. Seroconversion is not applicable to Day 1. Bars show mean values with 95% confidence intervals. Grey arrows indicate Dose 1 at Day 1 and Dose 2 at Day 29; immune assay blood draws preceded vaccination on each day. **Color key:** red = Group A (low dose), blue = Group B (medium dose), green = Group C (high dose), and brown = Group D (control).

### Rabies virus neutralizing antibody responses as measured by the Rapid Fluorescent Focus Inhibition Test (RFFIT)

At screening, eligibility required rabies virus neutralizing antibodies <0.5 IU/mL, below the WHO-defined seroprotection threshold. On Day 1, baseline GMTs were ≤0.3 IU/mL across all groups, and no measurable responses were detected by Day 8 (**Figure 4 and Supplemental Table 3**).

**FIGURE 4.**
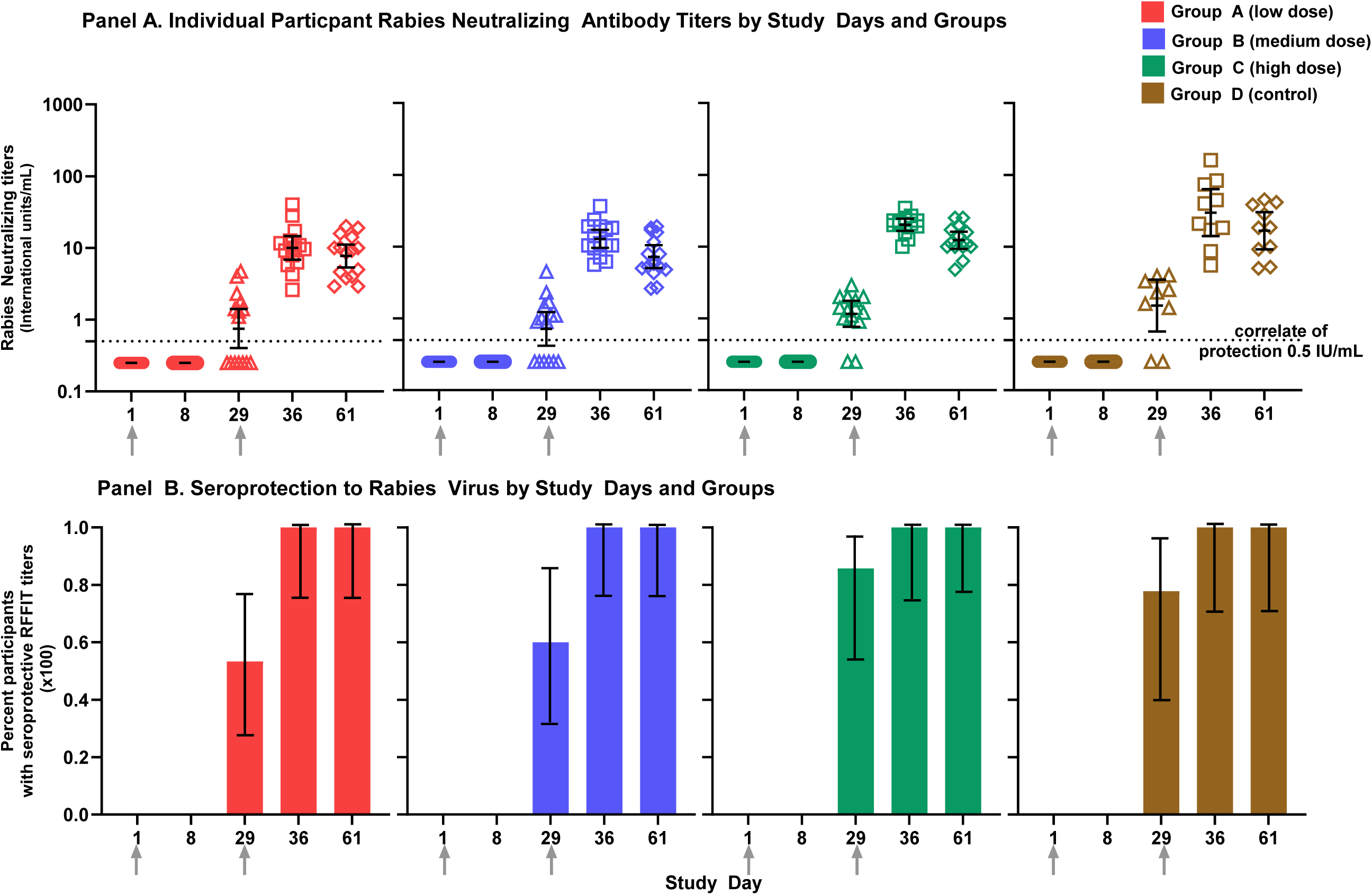
RABIES VIRUS ANTIBODY RESPONSES BY RAPID FLUORESCENT FOCUS INHIBITION TEST (RFFIT) TO RABIES VIRUS ACROSS STUDY GROUPS AND DAYS **Note:** **Panel A.** Rabies viral inhibition titers measured by rapid fluorescent focus inhibition test (RFFIT), shown by study arm and day. Individual symbols represent participant values; bars indicate geometric mean titers with 95 % confidence intervals (CIs). Grey arrows indicate Dose 1 at Day 1 and Dose 2 at Day 29; immune assay blood draws preceded vaccination on each day Symbols denote study day: solid circles = Day 0, open circles = Day 8, triangles = Day 29, squares = Day 36, and diamonds = Day 61. The dotted line represents the WHO-defined seroprotection threshold (≥0.5 IU/mL). **Panel B.** Proportion of participants in each study arm with seroprotection, defined as RFFIT value achieving the WHO correlate of protection. Bars show mean values with 95% confidence intervals. Grey arrows indicate Dose 1 at Day 1 and Dose 2 at Day 29; immune assay blood draws preceded vaccination on each day. **Color key**: red = Group A (low dose), blue = Group B (medium dose), green = Group C (high dose), and brown = Group D (control).

By Day 29, GMTs increased in all groups: 0.8 IU/mL (95% CI: 0.4–1.4) for Group A, 0.7 IU/mL (95% CI: 0.4–1.2) for Group B, 1.2 IU/mL (95% CI: 0.8–1.8) for Group C, and 1.5 IU/mL (95% CI: 0.7–3.5) for the control Group D. Seroprotection (RFFIT ≥0.5 IU/mL) was observed in 53.3%–85.7% of participants across groups. GMTs peaked on Day 36 and remained significantly above baseline, reaching 10.0 IU/mL (95% CI: 6.9–14.6) for Group A, 12.8 IU/mL (95% CI: 9.6–17.2) for Group B, 20.2 IU/mL (95% CI: 16.6–24.5) for Group C, and 29.7 IU/mL (95% CI: 14.0–63.2) for the control Group D. By Day 61, GMTs declined yet remained significantly above baseline: 7.7 IU/mL (95% CI: 5.3–11.1) in Group A, 7.2 IU/mL (95% CI: 5.0–10.5) in Group B, 12.3 IU/mL (95% CI: 9.3–16.2) in Group C, and 16.7 IU/mL (95% CI: 9.2–30.3) in the control Group D.

At Day 29, seroprotection was achieved in 53.3% (95% CI: 26.6–78.7) of Group A, 60.0% (95% CI: 32.3–83.7) of Group B, 85.7% (95% CI: 57.2–98.2) of Group C, and 77.8% (95% CI: 40.0–97.2) of the control Group D. From Day 36 through Day 61, after the second vaccination, seroprotection was 100% across all groups.

On Days 29 and 61, no statistically significant differences were observed across Groups A, B, C, and the control Group D, indicating comparable responses across all regimens and no optimal LASSARAB dose to advance for either a single- or two-dose strategy.

In preclinical studies, antibody responses measured by rabies virus glycoprotein IgG ELISA and by the RFFIT tended to align closely.^20–22^ To evaluate whether a similar relationship exists in humans, we compared antibody levels measured by both assays in the same participants and at corresponding study time points. ELISA and RFFIT titers were strongly associated (Spearman’s p = 0.89; 95% CI, 0.86–0.91; p < 0.001; n = 268), demonstrating a robust relationship between binding antibody concentrations and neutralizing activity.

### Safety

Among participants receiving the first vaccination with any product, immediate (within 30 minutes) reactogenicity symptoms were uncommon, all mild (Grade 1), and primarily related to local injection site reactions. Symptoms occurring in >10% of participants within seven days of the first vaccination with a LASSARAB-containing vaccine (n=44) included injection site tenderness (72.7%), pain (68.2%), malaise (43.2%), myalgia (38.6%), injection site warmth (29.5%), headache (25.0%), and feverishness (20.5%) (**Figure 5** and **Supplemental Table 4**). In the control Group D (n=10), symptoms occurring in >10% included injection site tenderness (60.0%), injection site pain (60.0%), myalgia (50.0%), headache (30.0%), malaise (30.0%), arthralgia (20.0%), and injection site warmth (20.0%). Reactogenicity across all four study groups was qualitatively similar. All solicited events were self-limited, mostly mild, and without severe (Grade 3) or greater events.

**FIGURE 5.**
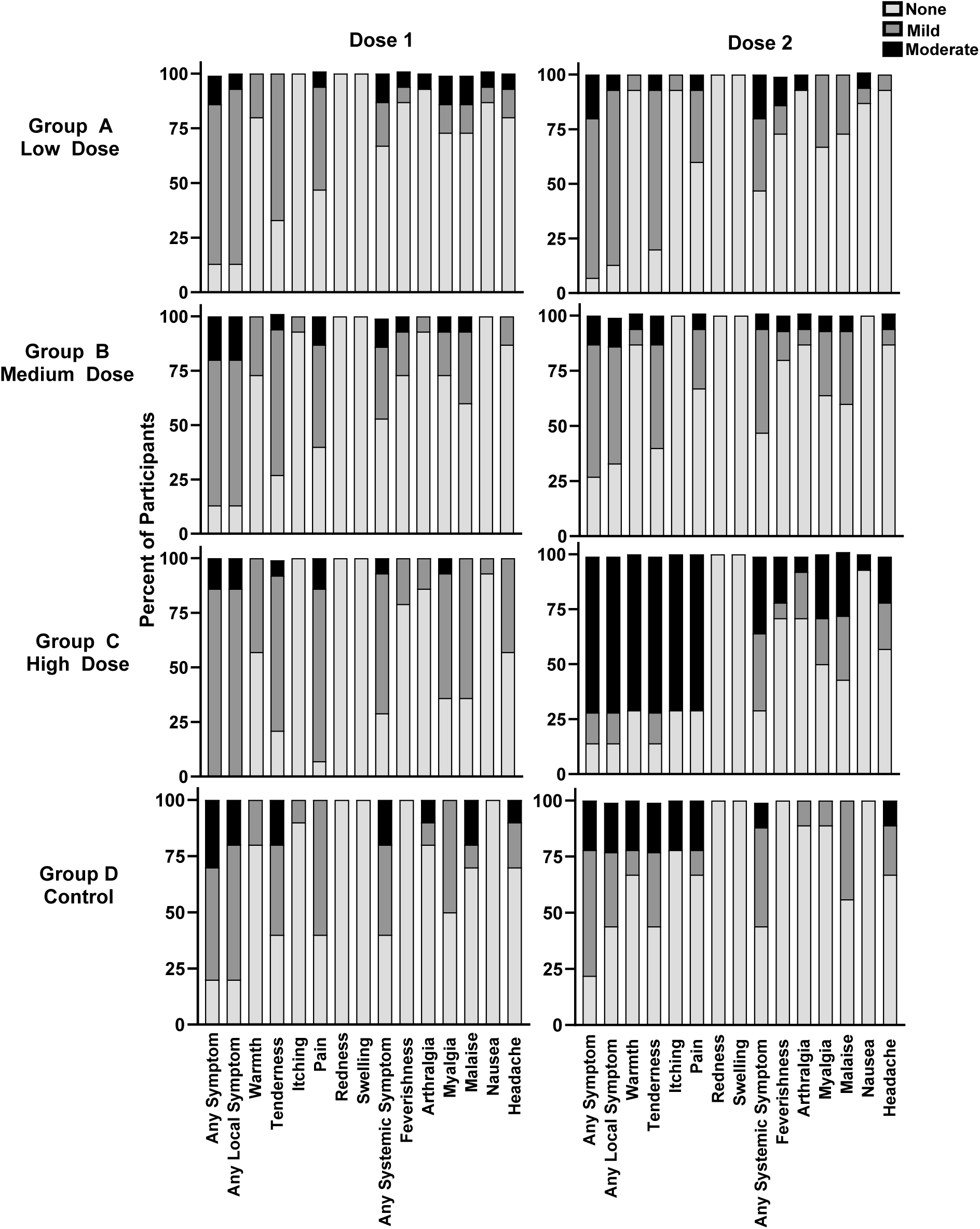
SOLICITED REACTOGENICITY THROUGH SEVEN DAYS AFTER VACCINATION BY GROUP **Note:** Solicited symptoms included local reactions (warmth, tenderness, itching, pain, redness, swelling) and systemic reactions (feverishness, arthralgia, myalgia, malaise, nausea, and headache). All participants who received a particular vaccine dose contributed safety data. The Dose 1 column shows the percentage of participants reporting solicited symptoms starting on Day 1, assessed within 30 minutes after vaccination and then daily through Day 8. The Dose 2 column shows the percentage of participants reporting solicited symptoms starting on Day 29, assessed within 30 minutes after vaccination and then daily through Day 36. Colors indicate the percentage of participants experiencing each severity grade: light grey = none (Grade 0), dark grey = mild (Grade 1), and black = moderate (Grade 2). No solicited adverse events of Grade 3 or higher were observed.

Among participants receiving the second vaccination, immediate (within 30 minutes) reactogenicity symptoms were also uncommon, all mild (Grade 1), and primarily injection-site related. Symptoms occurring in >10% of participants within seven days of a LASSARAB-containing vaccine (n=44) included injection site tenderness (75.0%), injection site pain (47.7%), malaise (40.9%), myalgia (38.6%), injection site warmth (29.5%), feverishness (25.0%), headache (20.5%), and arthralgia (15.9%) (**Figure 5** and **Supplemental Table 5**). In the control Group D (n=9), symptoms occurring in >10% included injection site tenderness (55.6%), malaise (44.4%), headache (33.3%), injection site pain (33.3%), injection site warmth (33.3%), arthralgia (11.1%), and myalgia (11.1%). Local reactogenicity was qualitatively higher in Group C compared with the other groups, while the remaining study groups had similar profiles. As with the first vaccination, events were self-limited, mostly mild, and without severe (Grade 3) or greater events.

There were five related unsolicited adverse events (AEs); two were of moderate severity and three were mild. All occurred within eight days of vaccination (four after the first dose and one after the second). There were no severe (Grade 3) or greater unsolicited AEs (**Supplemental Table 6**). No unsolicited event (related or unrelated) resulted in changes to the vaccination schedule. One related event and four unrelated events were unresolved by Day 61. There were five medically attended AEs, each moderate in severity, and none related to study vaccination.

There were no serious adverse events (SAEs), new-onset chronic medical conditions (NOCMCs), potentially immune mediated medical conditions (PIMMCs), or sensorineural hearing loss through Day 61. However, one unsolicited event was miscoded as a NOCMC and was not corrected before the Day 61 data freeze.

For this interim analysis, study investigators remain blinded to individual-level treatment group assignment. A line listing of blinded unsolicited AEs is presented in **Supplemental Table 7**. The final study report will include detailed descriptions of these events, including diagnosis and outcome, by treatment group through the end of the trial.

Clinical laboratory abnormalities were assessed through Day 61 (**Supplemental Table 6**). They were balanced across study groups. Of 20 abnormalities, 14 were mild and six were moderate. Eleven were considered to be related to the study vaccine. Of these, six were mild and five were moderate. There were no laboratory abnormalities of severe (Grade 3) or greater severity.

## DISCUSSION

This interim analysis of LASSARAB, a first-in-human, inactivated rabies-vectored Lassa virus GPC vaccine adjuvanted with a toll-like receptor-4–activating oil-in-water emulsion adjuvant (aPHAD-SE), demonstrated a favorable initial safety profile and 100% seroconversion as defined by ≥4-fold antibody rise against both Lassa and rabies viruses through 61 days of follow-up. Two intramuscular doses induced rapid and near-universal Lassa virus antibody seroconversion and achieved high rabies virus neutralizing antibody titers that met the established WHO correlate of protection.^23^ Reactogenicity was limited to transient, mostly mild local and systemic symptoms, and no SAEs or Grade 3 or greater AEs were observed. These findings support the continued development of this non-replicating Lassa fever vaccine candidate and underscore the potential of the rabies-vector platform to address the urgent need for countermeasures against two priority pathogens.

The preclinical evaluation of LASSARAB demonstrated strong immune responses against Lassa and rabies viruses in mouse and guinea pig models.^20^ Those results indicated that protection from Lassa virus was due, at least in part, to non-neutralizing, antibody-dependent, cellular functions. A subsequent study in cynomolgus macaques provided a foundation for our clinical trial observations.^21^ Two intramuscular doses adjuvanted with a TLR-4 agonist similar to that used in our study, aPHAD-SE, and administered 28 days apart elicited durable humoral responses to both the Lassa GPC and the rabies glycoprotein, with robust antibody titers persisting for more than one year. Following lethal Lassa virus challenge, all vaccinated primates survived and were protected from severe disease, while controls succumbed.^21^ Protection occurred despite the absence of pre-challenge Lassa virus neutralizing antibodies. The nonhuman primates vaccinated with LASSARAB developed non-neutralizing antibodies with Fc-γ receptor-mediated effector functions, as demonstrated by in vitro assays of antibody-dependent cellular cytotoxicity. Additionally, all vaccinated nonhuman primates achieved rabies virus-neutralizing antibody levels well above the WHO-defined correlate of protection. Our Phase 1 findings parallel these features: participants rapidly generated high titers of Lassa GPC-specific antibodies and attained rabies virus-neutralizing titers exceeding the same protective threshold. LASSARAB recipients in the highest dose group in this trial developed Lassa GPC-specific antibody titers in a similar range to those observed in the nonhuman primate model after vaccination. Concordance of these dual immune responses in both nonhuman primates and humans strengthens confidence in the rabies-vectored platform and its potential for dual pathogen protection.

Our findings are broadly consistent with the emerging experience from other Lassa virus vaccine candidates. Studies of a recombinant vesicular stomatitis virus (rVSV)-based Lassa virus vaccine candidate and a measles-vectored Lassa vaccine candidate have shown favorable safety profiles and rapid induction of Lassa virus-GPC antibodies.^24,25^ The robust Lassa virus GPC-specific antibody responses we observed align with these prior reports and reinforce the concept that humoral immunity to Lassa virus GPC can be achieved across diverse vaccine platforms.

The recent studies of rVSV- and measles-vectored vaccines illustrate both the promise and the distinctive characteristics of alternative Lassa fever vaccination strategies. The replication-competent rVSV-based Lassa virus vaccine has demonstrated efficacy in a nonhuman primate challenge study, with a single dose inducing strong Lassa virus GPC-specific binding and neutralizing antibodies, along with robust T-cell responses.^26–28^ One Phase 1 trial has been completed, and a Phase 2 trial is underway with this vaccine,^25,29,30^ but as of September 2025, human immune response data are not yet available to validate the nonhuman primate findings. While the only licensed vaccine using this platform, the rVSV-ZEBOV Ebola vaccine, is highly effective, it is not licensed for use in pregnant women, and its product insert advises caution in immunocompromised persons.^31^ Additionally, the Ebola vaccine requires ultra-cold storage (-80° C to -60° C),^31^ and large-scale rVSV vaccine production can be technically demanding for regional manufacturers.^32^ The measles-vectored Lassa vaccine also uses a live-attenuated, replication-competent backbone. In nonhuman primates and early-phase human studies, a single dose elicited Lassa-specific binding antibodies and T-cell responses.^33^ Similar to the rabies-vectored platform, neutralizing Lassa virus antibodies were low or absent after vaccination.^24,33^ The underlying measles vaccine platform is generally well tolerated in children and adults, but as a live virus vaccine, it is not recommended for routine use during pregnancy or in individuals with severe immunosuppression. Measles vaccine manufacturing capacity is well established worldwide, and lyophilized measles vaccines remain stable for years at standard cold-chain temperatures (2°C to 8°C).^34,35^

The rabies virus platform provides a well-established foundation for a dual-target vaccine.^36^ Since the early 1960s, inactivated rabies vaccines have been administered to millions globally, establishing a long and well-documented record of safety and efficacy,^37^ including in pregnant and immunocompromised persons.^23^ Moreover, licensed rabies vaccines are produced at global scale and can be lyophilized for thermostable storage,^38,39^ enabling distribution through existing manufacturing capacities and standard cold-chain systems. Building on this platform, the non-replicating rabies-vectored Lassa vaccine evaluated in our trial safely induced immune responses to both Lassa and rabies viruses in humans after two doses. In addition, a previous study showed that a similar rabies virus-based vaccine for Ebola virus can be heat-stabilized and stored for at least a year at room temperature.^40^ These data support the feasibility of a routine immunization strategy for regions where both pathogens are prevalent and cold-chain capacity is limited.

Rabies remains a major public health and veterinary challenge in many of the same West African countries where Lassa virus is endemic.^41,42^ In Nigeria, for example, the WHO estimates approximately 10,000 human deaths annually due to rabies--likely an undercount given very limited surveillance, especially in rural settings.^43,44^ Rabies control is further hindered by low vaccination coverage in dogs, shortages of human vaccines, weak infrastructure for post-exposure prophylaxis, and poor awareness of risk among both community and animal health sectors. The overlap in geography, risk, and health system constraints within Lassa endemic regions strengthens the rationale for a combination vaccine that addresses two urgent needs in a single intervention.

Rabies poses a unique challenge for vaccine development because of its nearly 100% case-fatality rate after symptom onset.^36^ Unlike diseases in which partial efficacy or herd immunity can reduce individual risk, rabies prevention requires complete and reliable protection in every vaccinated individual.^44^ Regulatory agencies and WHO guidelines mandate that rabies vaccine candidates elicit neutralizing antibody titers of ≥0.5 IU/mL in essentially all recipients following primary immunization, as this threshold remains the only validated correlate of protection.^23^ The rabies vaccine used for the control Group D is licensed for pre-exposure prophylaxis as either a two-dose regimen, administered on Days 0 and 7, or a three-dose regimen including an additional dose on Day 21 or 28.^45^ Rabies vaccine-induced immunity has been shown to persist for at least 10 years.^46^ In this Phase 1 trial, a single dose of the inactivated rabies-vectored Lassa vaccine achieved rabies virus seroprotection rates of up to 85.7%, while two doses induced 100% seroconversion. Across all time points, responses to LASSARAB-containing vaccines were statistically comparable to those elicited by the licensed rabies vaccine control.

Our trial has notable strengths. Combination vaccines have been identified as a global priority by the WHO.^47^ We implemented rigorous safety monitoring, including the use of Brighton Collaboration protocols for assessing sensorineural hearing loss,^48^ which provide high confidence in the observed tolerability profile. Immunogenicity was assessed with standardized assays, allowing direct comparison with prior nonhuman primate challenge data and with the established rabies correlate of protection.^23^ The strong concordance between rabies virus glycoprotein IgG ELISA and RFFIT titers suggests that ELISA may serve as a reliable surrogate for RFFIT in assessing immune responses to rabies-vectored vaccines, thereby streamlining future vaccine development. Finally, this study provides the first clinical evidence that a rabies-vectored vaccine can elicit concurrent immunity to two priority pathogens within a single product.

Our study has several limitations. As a Phase 1 trial, there were no pre-specified hypotheses to be tested, and the sample size was not chosen to detect group differences with sufficient statistical power. Next, enrollment was limited to healthy young and middle-aged adults in a region in which neither Lassa fever nor rabies is prevalent. This report includes safety and immunogenicity follow-up through Day 61. Longer-term data, including antibody durability and broader immunoprofiling, will follow trial completion and will provide a more complete picture of the vaccine’s immunological profile. As part of the clinical development pathway, trials in regions endemic for Lassa virus are in planning. Larger field studies will also be needed to define dosing, demonstrate protection, and identify correlates of protection.

In summary, this study provides the first clinical evidence that LASSARAB+aPHAD-SE, an adjuvanted inactivated rabies-vectored Lassa virus GPC vaccine, is well tolerated and elicits robust dual humoral immunity to Lassa and rabies viruses. Within 61 days, participants achieved Lassa seroconversion and rabies virus-neutralizing antibody titers exceeding the correlate of protection. Importantly, the safety profile was comparable to that of a licensed rabies vaccine. Together with the concordant protective efficacy shown in nonhuman primates, these data provide strong justification for advancing LASSARAB+aPHAD-SE into larger, longer-term trials in endemic settings and across diverse population groups. If efficacy is confirmed, this combination vaccine could help protect populations from two priority pathogens and deliver meaningful public health impact in regions where both diseases remain significant threats.

## ONLINE METHODS

### Overview

This study was conducted at the University of Maryland School of Medicine’s Center for Vaccine Development and Global Health (Baltimore, MD, USA). We enrolled healthy, non-pregnant, non-lactating adults 18 through 50 years of age. Participants were identified through the site’s volunteer database and by advertising in the Baltimore/Washington, DC area. Eligibility required completion of all screening assessments, including laboratory tests, pregnancy tests when indicated, rabies serology, audiometry, medical history, and physical examination. Individuals with clinically significant medical conditions or those taking medications that could affect safety or study outcomes were excluded.

There were four treatment groups (**Table 1 and Figure 1**). LASSARAB+aPHAD-SE was administered in three dose groups: Group A received 700 relative units (rU) of antigen with 5 µg of adjuvant in 1 mL, Group B received 1400 rU of antigen with 5 µg of adjuvant in 1 mL, and Group C received two concurrent doses of the medium-dose preparation for a total of 2800 rU of antigen and 10 µg of adjuvant. Group D, the control group, received the licensed inactivated rabies vaccine (Imovax Rabies; Sanofi Pasteur, Lyon, France) in its commercial presentation. Vaccine antigen content was measured by ELISA using a Lassa virus GPC recombinant protein as a standard and expressed as relative units (rUs), where 700rU of LASSA-GPC ELISA value is equivalent to approximately 50 µg of total virion protein content.

The LASSARAB-containing vaccines were prepared by the investigational pharmacy before administration. The final preparation appeared cloudy white. In contrast, the rabies vaccine comparator required reconstitution immediately prior to vaccination and appeared reddish-pink. To preserve blinding, unblinded vaccinators reconstituted the HDCV and drew it into the same type of needle and syringe used for the LASSARAB-containing vaccines, after which all syringes were masked with blue tape to obscure product color. Unblinded staff were responsible for vaccine accountability, storage, preparation, and administration, but were not involved in subsequent safety or immunogenicity assessments. Participants and all other study personnel were blinded to vaccine assignment and will continue to be blinded until study completion and database lock.

Eligible participants were enrolled and randomized to treatment groups using block randomization generated by the unblinded study statistician. Vaccinations were given on Days 1 and 29 by intramuscular injection in the preferred deltoid. For participants receiving two concurrent injections, bilateral deltoids were used. Participants were followed for vaccine safety and immunogenicity endpoints. The final study day is planned for Day 394. The study protocol defined an interim analysis of primary and secondary endpoints through Day 61. The primary objective of the study was to evaluate the safety and reactogenicity of LASSARAB+aPHAD-SE. Secondary objectives included assessments of Lassa and rabies virus immunogenicity on Days 1, 8, 29, 36, and 61.

### Study vaccines

#### Investigational vaccines

The study vaccine was a rabies-vectored, monovalent Lassa virus vaccine (LASSARAB) formulated with a synthetic 3D-(6-acyl) phosphorylated hexaacyl disaccharide (PHAD)-based stable squalene oil-in-water nanoemulsion adjuvant. LASSARAB was manufactured by IDT Biologika (Dessau-Rosslau, Germany; lot CTM0041222). The antigen consists of a chemically inactivated, genetically modified rabies virus vector engineered to express both the rabies virus glycoprotein and the full Lassa virus GPC.

The parental rabies virus backbone (BNSP) is a recombinant version of the SAD B19 strain, which has been widely used as a live oral rabies vaccine for wildlife in Europe.^49^ The relative rabies antigen content of LASSARAB compared to HDCV is not known. LASSARAB incorporates an arginine-to-glutamate (Arg→Glu) substitution at amino acid position 333 of the rabies glycoprotein, a modification known to reduce viral neurotropism.^50^ Following replication, viral particles are chemically inactivated with beta-propiolactone.

The adjuvant (aPHAD-SE) was produced by Curia Ltd. (Glasgow, UK; lot P07223/1). It is composed of a fully synthetic monophosphoryl lipid A (MPLA) analog, 3D-(6-acyl)-PHAD, combined with a stabilized squalene oil-in-water nanoemulsion (final squalene concentration 2% v/v). For administration, LASSARAB and aPHAD-SE were mixed immediately before use to yield a single 1-mL intramuscular dose, given as a two-dose series 28 days apart.

### Control Vaccine

The control vaccine was the licensed human diploid cell vaccine (HDCV)--sold as Imovax Rabies and produced by Sanofi Pasteur (Lyon, France). HDCV is a sterile, stable, freeze-dried suspension of rabies virus (strain PM-1503-3M, derived from the Pitman-Moore lineage) grown in MRC-5 human diploid cells, concentrated by ultrafiltration, and inactivated with beta-propiolactone.^45^ The vaccine is provided in single-dose vials without preservatives. Each 1 mL dose contains ≥2.5 international units (IU) of rabies antigen.^45^ HDCV is indicated for pre- and post-exposure prophylaxis and approved for use in all age groups.^45^ HDCV is licensed for pre-exposure prophylaxis as either a two-dose regimen, administered on Days 0 and 7, or a three-dose regimen with a third dose on Day 21 or 28.^45^ A Rapid Fluorescent Focus Inhibition Test (RFFIT) titer ≥0.5 IU/mL is considered seroprotective.^51^

### Normal Saline Placebo

Sterile 0.9% sodium chloride for injection (USP) was administered as an intramuscular injection concurrent with HDCV in cohorts that included Group C. This maintained blinding, as Group C received two concurrent intramuscular injections.

### Procedures

Participants gave written informed consent before any data were collected or study procedures initiated. Eligible participants were enrolled and assigned to treatment groups using block randomization prepared by the unblinded study statistician. Participants were enrolled and randomized sequentially in a stepwise, dose-escalation process using four vaccination cohorts (**Supplemental Table 1**). Participants were blinded to vaccine allocation, as were study staff and investigators involved in subsequent assessments of trial endpoints. Only trial personnel involved in receipt, storage, preparation, and administration of the study product were unblinded.

If all participants proceeded to vaccination, the final vaccine assignment ratio would be 3:3:3:2 for Group A (low dose), Group B (medium dose), Group C (high dose), and the control Group D. Intramuscular injection was administered in the preferred deltoid (single vaccination groups) or bilateral deltoids (double vaccination groups). Vaccination occurred on Days 1 and 29. After each vaccination, participants were observed in the study clinic for 30 minutes. Participants had vital signs recorded, were examined by a study clinician (if indicated), completed an immediate reactogenicity assessment, received paper symptom diary training, and were discharged.

Two days after each vaccination, participants received a follow-up phone call to evaluate AEs, review diary instructions, and confirm their next appointment. They returned to the clinic on Days 8 and 36 for safety evaluations, during which a study investigator reviewed the paper diaries, assessed vaccine reactogenicity, and documented any other safety issues. Venous blood samples were collected at these visits for hematology and chemistry testing. Hearing thresholds to evaluate for new-onset sensorineural hearing loss, were assessed by a validated tablet-based audiometer at screening, after the first vaccine dose at Day 22, after the second vaccine dose at Day 61, upon completion of the study at Day 394, and at any time if the participant reported a change in hearing or if deemed necessary by study investigators. Additional safety and immunogenicity assessments were conducted on Days 36 and 61. Serum samples (15 mL) for vaccine immune response measurements were collected on Days 1, 8, 29, 36, and 61.

After seven days of safety data had been collected for each of the first three cohorts, the Safety Review Committee met to evaluate the findings. Following predefined criteria, the committee determined whether to continue vaccinating the remaining participants at the current dose level and to initiate enrollment of additional participants at the next higher dose.

Study data were collected and managed using REDCap electronic data capture tools hosted at the University of Maryland School of Medicine.^52,53^ REDCap (Research Electronic Data Capture) is a secure, web-based software platform designed to support data capture for research studies, providing (1) an intuitive interface for validated data capture; (2) audit trails for tracking data manipulation and export procedures; (3) automated export procedures for seamless data downloads to common statistical packages; and (4) procedures for data integration and interoperability with external sources.

### Safety assessments

Vaccine reactogenicity was evaluated by monitoring solicited local and systemic reactions from the time of each vaccination through seven days post-vaccination. Local reactions included warmth, tenderness, itching, pain, redness, and swelling, while systemic reactions included feverishness, arthralgia, myalgia, malaise, nausea, and headache.

We assessed unsolicited AEs, SAEs, MAAEs, NOCMCs, PIMMCs, and an AE of special interest (AESI) from enrollment through Day 61. We also assessed clinical laboratory abnormalities through Day 61.

Given theoretical risks of sensorineural hearing loss after Lassa virus vaccination,^48^ we predefined sensorineural hearing loss as an AESI. Evaluation of sensorineural hearing loss followed Brighton Collaboration guidance.^48^ Participants underwent scheduled evaluations for sensorineural hearing loss at eligibility assessment, at three scheduled time points after vaccination, and at unscheduled assessments if the participant reported a change in hearing or at the discretion of the investigator. To follow up on abnormal in-clinic audiology tests after enrollment, participants were referred for formal diagnostic sound booth audiometry completed by a licensed audiologist and evaluation by an otologist for sensorineural hearing loss confirmation. During eligibility assessment, we excluded individuals with a baseline hearing level >15 dB at three consecutive frequencies or ≥20 dB at any single frequency by tablet audiometry, with exam confirmation of sensorineural hearing loss.

### Planned assessments after this interim analysis

The study protocol provides for participant safety monitoring through study Day 394 (approximately one year after the second study vaccination). Additional safety and immunogenicity visits are scheduled for Days 121 and 394. A safety phone call is scheduled for Day 210. Additional protocol-defined humoral immunogenicity assessments are scheduled for Days 121 and 394. Audiometry is scheduled for Day 394. SAEs, MAAEs, NOCMCs, PIMMCs, and AESIs will be monitored through Day 394.

Further exploratory immunogenicity analyses will be conducted post-trial using clinical specimens collected for future use.

### Immunogenicity assessments

We assessed immune responses to Lassa and rabies viruses using multiple assays at various time points:

- Lassa virus GPC antibodies by ELISA at Days 1, 8, 29, 36, and 61
- Rabies virus glycoprotein antibodies by ELISA at Days 1, 8, 29, 36, and 61
- Rabies viral neutralization by Rapid Fluorescent Focus Inhibition Test (RFFIT) at Days 1, 8, 29, 36, and 61

### Lassa virus GPC antibodies and rabies virus glycoprotein antibodies

ELISAs developed for non-human primates were adapted and qualified to quantify Lassa virus- and rabies virus-specific human IgG.^22^ The qualified assays incorporated the use of the First WHO International Standard for anti-Lassa virus antibodies (NIBSC Code 20/202, Potters Bar, UK) and an in-house standard calibrated against the Third WHO International Standard for anti-rabies immunoglobulin (NIBSC Code 19/244, Potters Bar, UK).

Immulon 2HB 96-well microplates were coated with recombinant Lassa virus GPC or rabies virus glycoprotein produced at Thomas Jefferson University at 0.5 μg/mL in bicarbonate buffer and incubated for 16 to 18 hours at 4°C. Plates were washed using a BioTek 405 TS microplate washer (BioTek, Santa Clara, CA) and blocked with 5% non-fat dry milk (NFDM) in phosphate-buffered saline (PBS) containing 0.05% Tween-20 (PBS-TM) for 1 hour at room temperature (22°C).

Following blocking, serum samples and controls diluted in PBS-TM were added in duplicate and incubated for 2 hours at room temperature. After washing, bound antibodies were detected using HRP-conjugated goat anti-human IgG (Jackson ImmunoResearch, West Grove, PA), followed by TMB substrate. The reaction was stopped after 15 minutes with 1 M phosphoric acid, and absorbance at 450 nm was measured using an AccuSkan FC microplate photometer with Skanlt Software (Thermo Fisher Scientific, Waltham, MA).

Each assay included an 8-point calibration curve of the corresponding standard. Antibody concentrations (IU/ml) were determined by interpolation from the calibration curve using GraphPad Prism version 10 (GraphPad, Boston, MA) and adjusted for sample dilution.

### Rabies virus antibodies by Rapid Fluorescent Foci Inhibition Test (RFFIT) test

The RFFIT was conducted as previously described.^54^ Human serum samples were serially diluted 2-fold in 96-well plates using Opti-MEM medium (Invitrogen, Waltham, MA), starting at a 1:4 dilution for the prescreen and a 1:5 dilution for immunized participants. Each plate included a serial dilution of the Third WHO International Standard for Anti-Rabies Immunoglobulin, SRIG, NIBSC code: 19/244, in duplicates, beginning at 0.5 IU/ml, to create a calibration curve.

A volume of rabies virus strain CVS-11, previously determined to infect 90% of confluent cells, was added to each well containing the diluted sera/SRIG and incubated for 1 hour at 34°C. Subsequently, BSR cells (a clone of baby hamster kidney cells; 6 × 10^4^ per well) were added to the serum/virus mixture, and plates were incubated at 34°C for an additional 23 hours. Cells were then fixed with 80% acetone and stained with FITC-conjugated anti-rabies virus nucleoprotein antibody.

The percentage of infection was evaluated using fluorescence microscopy and a Cytation 5 reader (Agilent BioTek, Santa Clara, CA). Fifty percent endpoint titers were calculated by the Reed-Muench method and converted to IU/mL by comparison with the SRIG calibration curve. Samples that did not reach a 50% endpoint were reanalyzed at a higher starting dilution.^55^

### Statistical analysis

Analyses were descriptive; no confirmatory hypothesis testing was prespecified. The planned sample size (N=55: 15 per experimental vaccine group; 10 HDCV controls) was chosen to characterize safety and immunogenicity, not to power between-group comparisons, and was consistent with FDA guidance.^56^

All participants who were randomized and received at least an initial dose of study vaccine were included in the safety analysis. For the immunogenicity analysis, all participants who received at least an initial dose of study vaccine and for whom data were available at a particular time point were included. Outcomes were analyzed by vaccine group. Baseline demographics were summarized descriptively. Safety endpoints were tabulated by severity grade and relationship to vaccination. Serum antibodies were described in ELISA units or RFFIT titers. Seroconversion for ELISA assays was defined as a ≥4-fold rise from the Day 1 baseline. Seroprotection for RFFIT was defined as titers ≥0.5 IU/mL, consistent with WHO guidance.^23^ ELISA data were summarized by GMT, GMFR from Day 1, and the proportion of participants achieving seroconversion. Missing data were not imputed for safety or immunogenicity analyses.

For binary outcomes such as ELISA seroconversion (Yes/No) or RFFIT seroprotection (Yes/No), proportions were calculated for each vaccine group with 95% confidence intervals using the Clopper-Pearson exact method. Differences in continuous outcomes between the four vaccine groups were assessed using the Kruskal-Wallis H-test, with pairwise comparisons conducted. Within-group changes in antibody titers across time points were evaluated using the Wilcoxon signed-rank test. Spearman’s rank correlation was computed to assess the relationship between rabies virus glycoprotein ELISA titers and RFFIT neutralization titers. No adjustments were made for multiple comparisons. All statistical tests were two-sided. Analyses were performed using SAS version 9.4 (SAS Institute, Cary, NC). Figures were generated using GraphPad Prism version 10 (GraphPad Software, Boston, MA).

### Regulatory and ethics

The protocol and informed consent forms were approved by the University of Maryland, Baltimore, Institutional Review Board (HP-00110576). The trial was registered at Clinicaltrials.gov (NCT06546709) on December 4, 2024. The trial protocol and statistical analysis plan will be uploaded to Clinicaltrials.gov upon study completion.

## Data Availability

The data that support the findings of this study are available from the corresponding author upon reasonable request after study completion and study database lock.

## ACKNOWLEDGEMENTS

We thank our study volunteers, and the administrative, clinical, laboratory, recruiting, and regulatory staff of the Center for Vaccine Development and Global Health at the University of Maryland School of Medicine. We also thank the administrative and laboratory staff of the Department of Microbiology and Immunology at Thomas Jefferson University. We thank Dr. Maryam Jahromi for serving as a member of the trial Safety Review Committee and Dr. Kawsar Talaat for serving as the trial Independent Safety Monitor. We are grateful to our colleagues at the National Institute of Allergy and Infectious Diseases for technical advice.

The funders had no role in study design, data collection and analysis, decision to publish, or preparation of the manuscript. The content of this publication and any associated federal funding does not necessarily represent the views or policies of the U.S. Department of Health and Human Services, nor does mention of trade names, commercial products, or organizations imply endorsement by the U.S. Government. Opinions, interpretations, conclusions, and recommendations are those of the authors and do not necessarily reflect endorsement by the U.S. Government.

## ROLE OF THE FUNDING SOURCE

The National Institute of Allergy and Infectious Diseases (NIAID) funded the study and offered guidance on immunogenicity and safety assessments, but had no role in the design, conduct, analysis, interpretation, or writing of the manuscript.

Justin R. Ortiz had complete access to all study data, accepts responsibility for the integrity and accuracy of the analyses, and made the final decision to submit the manuscript for publication.

## PATIENT AND PUBLIC INVOLVEMENT

Patients or the public were not involved in the design, conduct, reporting, or dissemination plans of this research.

## DATA AVAILABILITY

The data that support the findings of this study are available from the corresponding author upon reasonable request. Reagents will be made available upon reasonable request. Data are in controlled access data storage at the University of Maryland School of Medicine.

## CODE AVAILABILITY

The study did not use custom code or mathematical algorithms to generate results.

