## Supplement for "Immunogenicity of an adjuvanted, combination inactivated rabies-vectored, Lassa fever vaccine in healthy adults: interim results of a first-in-human Phase 1 trial"

**ONLINE SUPPLEMENT**

### SUPPLEMENTAL TABLE 1. PROTOCOL DOSE ESCALATION SCHEDULE

| Cohort | Group | Number of Participants | Group Type | Injection #1 Treatment | Injection #2 Treatment |
|---|---|---|---|---|---|
| 1 | A | 4 | Sentinel | LASSARAB 700 rU Dose | N/A |
| | D | 1 | Control | HDCV control | N/A |
| | Total | 5 | | | |
| 2 | A | 11 | Expanded | LASSARAB 700 rU Dose | N/A |
| | B | 4 | Sentinel | LASSARAB 1400 rU Dose | N/A |
| | D | 3 | Control | HDCV control | N/A |
| | Total | 18 | | | |
| 3 | B | 11 | Expanded | LASSARAB 1400 rU Dose | N/A |
| | D | 2 | Control | HDCV control | N/A |
| | C | 4 | Sentinel | LASSARAB 1400 rU Dose | LASSARAB 1400 rU Dose |
| | D | 1 | Control | HDCV control | Normal saline placebo |
| | Total | 18 | | | |
| 4 | C | 11 | Expanded | LASSARAB 1400 rU Dose | LASSARAB 1400 rU Dose |
| | D | 3 | Control | HDCV control | Normal saline placebo |
| | Total | 14 | | | |

**Note:** Treatment Group C receives two vaccine injections administered in bilateral arms concurrently. To maintain blinding of participants in Cohorts 3 and 4, one dose of normal saline placebo is administered in the contralateral arm to one participant receiving HDCV control in Cohort 3 and to all three participants receiving HDCV control in Cohort 4.

### SUPPLEMENTAL TABLE 2. LASSA VIRUS ANTIBODY ELISA RESPONSES

| Treatment Group | Group A | Group A (post hoc) | Group B | Group C | Group D |
|---|---|---|---|---|---|
| **Description** | **Low Dose** | **Low Dose** | **Medium Dose** | **High Dose** | **Control** |
| Day 1, n= | 15 | 14 | 15 | 14 | 10 |
| GMT (95% CI) | 1.7 (0.8–3.6) | 1.2 (0.8–1.8) | 1.8 (1.0–3.0) | 2.1 (1.0–4.6) | 1.4 (0.7–2.8) |
| Day 8, n= | 15 | 14 | 15 | 14 | 10 |
| GMT (95% CI) | 2.0 (0.7–6.1) | 1.2 (0.8–2.0) | 1.7 (0.9–3.1) | 2.6 (1.4–4.8) | 1.6 (0.8–3.0) |
| %seroconversion (95% CI) | 6.7 (0.0–32.0) | 0.0 (0.0–23.2) | 0.0 (0.0–21.8) | 0.0 (0.0–23.2) | 0.0 (0.0–30.9) |
| GMFR (95% CI) | 1.2 (0.8–1.8) | 1.0 (0.8–1.2) | 0.9 (0.6–1.5) | 1.2 (0.8–1.8) | 1.1 (0.7–1.7) |
| Day 29, n= | 15 | 14 | 15 | 14 | 9 |
| GMT (95% CI) | 8.8 (3.3–23.6)* | 5.8 (3.5–9.7)* | 6.4 (4.0–10.3)* | 13.4 (9.6–18.9)*§ | 1.4 (0.7–2.7)# |
| %seroconversion (95% CI) | 60.0 (32.3–83.7) | 57.1 (28.9–82.3) | 33.3 (11.8–61.6) | 64.3 (35.1–87.2) | 0.0 (0.0–33.6) |
| GMFR (95% CI) | 5.2 (3.1–8.9) | 4.8 (2.8–8.1) | 3.6 (2.4–5.5) | 6.4 (3.0–13.6) | 1.0 (0.6–1.6) |
| Day 36, n= | 15 | 14 | 15 | 13 | 10 |
| GMT (95% CI) | 96.7 (45.7–204.3)* | 75.7 (42.5–134.9)* | 117.6 (53.7–257.6)* | 295.4 (196.9–443.0)*§ | 1.4 (0.7–2.8)# |
| %seroconversion (95% CI) | 100.0 (78.2–100.0) | 100.0 (76.8–100.0) | 93.3 (68.1–99.8) | 100.0 (75.3–100.0) | 0 (0.0–30.9) |
| GMFR (95% CI) | 57.7 (30.7–108.4) | 61.9 (31.9–120.0) | 66.8 (28.4–156.7) | 156.1 (68.2–357.1) | 1.0 (0.6–1.5) |
| Day 61, n= | 15 | 14 | 15 | 14 | 10 |
| GMT (95% CI) | 158.8 (81.1–310.9)* | 130.0 (74.3–227.4)* | 135.4 (87.6–209.3)* | 331.0 (174.0–629.6)* | 1.5 (0.7–3.2)# |
| %seroconversion (95% CI) | 100.0 (78.2–100.0) | 100.0 (76.8–100.0) | 100.0 (78.2–100.0) | 100.0 (76.8–100.0) | 0.0 (0.0–30.9) |
| GMFR (95% CI) | 94.8 (54.1–166.1) | 106.3 (61.7–183.4) | 76.9 (38.2–154.9) | 157.2 (67.8–364.6) | 1.0 (0.5–2.1) |

**Note:** *Wilcoxon signed-rank test for within-group comparison to baseline (Day 1), p <0.05. No dose response was observed among Groups A–C. One Group A participant had a high baseline Lassa antibody titer, so we performed a post hoc analysis excluding this individual. Lassa virus ELISA responses for this analysis are in “Group A (post hoc).” #Kruskal-Wallis Test Group D is significantly different than Group A (post hoc), B and C, p<0.05. §Kruskal-Wallis Test Group C is significantly different than Group A (post hoc), p<0.05. CI=confidence interval; GMT=geometric mean titer; seroconversion (ELISA) defined as a ≥4-fold rise from Day 1; GMFR=geometric mean fold rise.

### SUPPLEMENTAL TABLE 3. RABIES VIRUS ANTIBODY ELISA RESPONSES AND RAPID FLUORESCENT FOCI INHIBITION TEST

| Treatment Group | Group A | Group B | Group C | Group D |
|---|---|---|---|---|
| **Product** | **Low Dose** | **Medium Dose** | **High Dose** | **Control** |
| **ELISA** | | | | |
| Day 1, n= | 15 | 15 | 14 | 10 |
| GMT (95% CI) | 0.04 (0.03–0.06) | 0.02 (0.02–0.04) | 0.04 (0.02–0.07) | 0.02 (0.01–0.03) |
| Day 8, n= | 15 | 15 | 14 | 10 |
| GMT (95% CI) | 0.06 (0.04–0.09)* | 0.2 (0.1–0.3)* | 0.1 (0.05–0.2)* | 0.1 (0.03–0.3)* |
| %seroconversion (95% CI) | 0.0 (0.0–21.8) | 66.7 (38.4–88.2) | 28.6 (8.4–58.1) | 50.0 (18.7–81.3) |
| GMFR (95% CI) | 1.4 (1.1–1.9) | 7.3 (4.2–12.9) | 2.9 (1.5–5.7) | 6.4 (2.0–20.2) |
| Day 29, n= | 15 | 15 | 14 | 9 |
| GMT (95% CI) | 5.9 (3.5–9.8)* | 8.6 (5.8–12.8)* | 19.0 (12.6–28.7)*# | 1.7 (0.9–3.3)*# |
| %seroconversion (95% CI) | 100.0 (78.2–100.0) | 100.0 (78.2–100.0) | 100.0 (76.8–100.0) | 100.0 (66.4–100.0) |
| GMFR (95% CI) | 131.7 (71.2–243.5) | 356.9 (228.0–558.6) | 542.1 (248.8–1181.3) | 130.4 (55.8–304.5) |
| Day 36, n= | 15 | 15 | 13 | 10 |
| GMT (95% CI) | 45.0 (29.5–68.6)* | 73.2 (45.6–117.5)* | 170.3 (128.3–226.2)*# | 26.5 (11.8–59.5)* |
| %seroconversion (95% CI) | 100.0 (78.2–100.0) | 100.0 (78.2–100.0) | 100.0 (75.3–100.0) | 100.0 (69.2–100.0) |
| GMFR (95% CI) | 1011.9 (579.0–1768.4) | 3036.1 (1815.7–5076.9) | 5896.9 (3746.0–9282.8) | 1750.7 (502.3–6101.9) |
| Day 61, n= | 15 | 15 | 14 | 10 |
| GMT (95% CI) | 38.0 (24.4–59.3)* | 51.1 (35.9–72.9)* | 90.0 (63.3–127.9)*# | 14.6 (8.0–26.7)*# |
| %seroconversion (95% CI) | 100.0 (78.2–100.0) | 100.0 (78.2–100.0) | 100.0 (76.8–100.0) | 100.0 (69.2–100.0) |
| GMFR (95% CI) | 855.0 (499.2–1464.3) | 2121.9 (1438.0–3131.0) | 2565.6 (1164.1–5654.5) | 963.7 (339.9–2732.3) |
| **Rapid Fluorescent Foci Inhibition Test** | | | | |
| Day 1, n= | 15 | 15 | 14 | 10 |
| GMT (95% CI) | 0.3 (0.3–0.3) | 0.3 (0.3–0.3) | 0.3 (0.3–0.3) | 0.3 (0.3–0.3) |
| % seroprotected (95% CI) | 0.0 (0.0-21.8) | 0.0 (0.0-21.8) | 0.0 (0.0-23.2) | 0.0 (0.0-30.9) |
| Day 8, n= | 15 | 15 | 14 | 10 |
| GMT (95% CI) | 0.3 (0.3–0.3) | 0.3 (0.3–0.3) | 0.3 (0.3–0.3) | 0.3 (0.3–0.3) |
| % seroprotected (95% CI) | 0.0 (0.0-21.8) | 0.0 (0.0-21.8) | 0.0 (0.0-23.2) | 0.0 (0.0-30.9) |
| GMFR (95% CI) | 1.0 (1.0–1.0) | 1.0 (1.0–1.0) | 1.0 (1.0–1.0) | 1.0 (1.0–1.0) |
| Day 29, n= | 15 | 15 | 14 | 9 |
| GMT (95% CI) | 0.8 (0.4–1.4)* | 0.7 (0.4–1.2)* | 1.2 (0.8–1.8)* | 1.5 (0.7–3.5)* |
| % seroprotected (95% CI) | 53.3 (26.6–78.7) | 60.0 (32.3–83.7) | 85.7 (57.2–98.2) | 77.8 (40.0–97.2) |
| GMFR (95% CI) | 3.0 (1.6–5.6) | 2.9 (1.7–4.9) | 4.6 (3.1–7.0) | 6.1 (2.6–13.9) |
| Day 36, n= | 15 | 15 | 13 | 10 |
| GMT (95% CI) | 10.0 (6.9–14.6)* | 12.8 (9.6–17.2)* | 20.2 (16.6–24.5)*§ | 29.7 (14.0–63.2)* |
| % seroprotected (95% CI) | 100.0 (78.2–100.0) | 100.0 (78.2–100.0) | 100.0 (75.3–100.0) | 100.0 (69.2–100.0) |
| GMFR (95% CI) | 40.1 (27.5–58.4) | 51.3 (38.4–68.6) | 80.7 (66.4–98.0) | 118.9 (55.9–252.8) |
| Day 61, n= | 15 | 15 | 14 | 10 |
| GMT (95% CI) | 7.7 (5.3–11.1)* | 7.2 (5.0–10.5)* | 12.3 (9.3–16.2)* | 16.7 (9.2–30.3)* |
| % seroprotected (95% CI) | 100.0 (78.2–100.0) | 100.0 (78.2–100.0) | 100.0 (76.8–100.0) | 100.0 (69.2–100.0) |
| GMFR (95% CI) | 30.8 (21.3–44.5) | 29.0 (20.0–42.0) | 49.2 (37.3–64.9) | 66.7 (36.7–121.1) |

**Note:** Wilcoxon signed-rank test for within-group comparison to baseline (Day 1), p<0.05. #Kruskal–Wallis Test: Day 29—Group C is significantly different than Groups A and B; Group D is significantly different than Groups A, B, and C (p<0.05). Day 36—Group C is significantly different than Groups A, B, and D (p<0.05). Day 61—Group D is significantly different than Groups B and C; Group C is significantly different than Group A (p<0.05). §Kruskal–Wallis Test: Day 36—Group C is significantly different than Group A (p<0.05). CI=confidence interval; GMT=geometric mean titer; seroconversion (ELISA) defined as ≥4-fold rise from Day 1; GMFR=geometric mean fold rise.

### SUPPLEMENTAL TABLE 4. SOLICITED EVENTS AFTER THE FIRST VACCINE DOSE

| **Treatment Group** | **Group A n (%)** | **Group B n (%)** | **Group C n (%)** | **Group D n (%)** | **Any LASSARAB-containing vaccine** | **Overall n (%)** |
|---|---|---|---|---|---|---|
| **Product** | **Low Dose** | **Medium Dose** | **High Dose** | **Control** | | |
| Participants, n= | 15 | 15 | 14 | 10 | 44 | 54 |
| Any Symptom | 13 (86.7%) | 13 (86.7%) | 14 (100.0%) | 8 (80.0%) | 40 (90.9%) | 48 (88.9%) |
| Any Local Symptom | 13 (86.7%) | 13 (86.7%) | 14 (100.0%) | 8 (80.0%) | 40 (90.9%) | 48 (88.9%) |
| Warmth | 3 (20.0%) | 4 (26.7%) | 6 (42.9%) | 2 (20.0%) | 13 (29.5%) | 15 (27.8%) |
| Tenderness | 10 (66.7%) | 11 (73.3%) | 11 (78.6%) | 6 (60.0%) | 32 (72.7%) | 38 (70.4%) |
| Itching | 0 (0.0%) | 1 (6.7%) | 0 (0.0%) | 1 (10.0%) | 1 (2.3%) | 2 (3.7%) |
| Pain | 8 (53.3%) | 9 (60.0%) | 13 (92.9%) | 6 (60.0%) | 30 (68.2%) | 36 (66.7%) |
| Redness | 0 (0.0%) | 0 (0.0%) | 0 (0.0%) | 0 (0.0%) | 0 (0.0%) | 0 (0.0%) |
| Swelling | 0 (0.0%) | 0 (0.0%) | 0 (0.0%) | 0 (0.0%) | 0 (0.0%) | 0 (0.0%) |
| Any Systemic Symptom | 5 (33.3%) | 7 (46.7%) | 10 (71.4%) | 6 (60.0%) | 22 (50.0%) | 28 (51.9%) |
| Feverishness | 2 (13.3%) | 4 (26.7%) | 3 (21.4%) | 0 (0.0%) | 9 (20.5%) | 9 (16.7%) |
| Arthralgia | 1 (6.7%) | 1 (6.7%) | 2 (14.3%) | 2 (20.0%) | 4 (9.1%) | 6 (11.1%) |
| Myalgia | 4 (26.7%) | 4 (26.7%) | 9 (64.3%) | 5 (50.0%) | 17 (38.6%) | 22 (40.7%) |
| Malaise | 4 (26.7%) | 6 (40.0%) | 9 (64.3%) | 3 (30.0%) | 19 (43.2%) | 22 (40.7%) |
| Nausea | 2 (13.3%) | 0 (0.0%) | 1 (7.1%) | 0 (0.0%) | 3 (6.8%) | 3 (5.6%) |
| Headache | 3 (20.0%) | 2 (13.3%) | 6 (42.9%) | 3 (30.0%) | 11 (25.0%) | 14 (25.9%) |

**Note:** Presented data are n (%) of participants experiencing each solicited symptom from vaccination through Day 8.

### SUPPLEMENTAL TABLE 5. SOLICITED EVENTS AFTER THE SECOND VACCINE DOSE

| Treatment Group | Group A n (%) | Group B n (%) | Group C n (%) | Group D n (%) | Any LASSARAB-containing vaccine | Overall n (%) |
|---|---|---|---|---|---|---|
| **Product** | **Low Dose** | **Medium Dose** | **High Dose** | **Control** | | |
| Participants, n= | 15 | 15 | 14 | 9 | 44 | 53 |
| Any Symptom | 14 (93.3%) | 11 (73.3%) | 12 (85.7%) | 7 (77.8%) | 37 (84.1%) | 44 (83.0%) |
| Any Local Symptom | 13 (86.7%) | 10 (66.7%) | 12 (85.7%) | 5 (55.6%) | 35 (79.5%) | 40 (75.5%) |
| Warmth | 1 (6.7%) | 2 (13.3%) | 10 (71.4%) | 3 (33.3%) | 13 (29.5%) | 16 (30.2%) |
| Tenderness | 12 (80.0%) | 9 (60.0%) | 12 (85.7%) | 5 (55.6%) | 33 (75.0%) | 38 (71.7%) |
| Itching | 1 (6.7%) | 0 (0.0%) | 10 (71.4%) | 2 (22.2%) | 11 (25.0%) | 13 (24.5%) |
| Pain | 6 (40.0%) | 5 (33.3%) | 10 (71.4%) | 3 (33.3%) | 21 (47.7%) | 24 (45.3%) |
| Redness | 0 (0.0%) | 0 (0.0%) | 0 (0.0%) | 0 (0.0%) | 0 (0.0%) | 0 (0.0%) |
| Swelling | 0 (0.0%) | 0 (0.0%) | 0 (0.0%) | 0 (0.0%) | 0 (0.0%) | 0 (0.0%) |
| Any Systemic Symptom | 8 (53.3%) | 8 (53.3%) | 10 (71.4%) | 5 (55.6%) | 26 (59.1%) | 31 (58.5%) |
| Feverishness | 4 (26.7%) | 3 (20.0%) | 4 (28.6%) | 0 (0.0%) | 11 (25.0%) | 11 (20.8%) |
| Arthralgia | 1 (6.7%) | 2 (13.3%) | 4 (28.6%) | 1 (11.1%) | 7 (15.9%) | 8 (15.1%) |
| Myalgia | 5 (33.3%) | 5 (33.3%) | 7 (50.0%) | 1 (11.1%) | 17 (38.6%) | 18 (34.0%) |
| Malaise | 4 (26.7%) | 6 (40.0%) | 8 (57.1%) | 4 (44.4%) | 18 (40.9%) | 22 (41.5%) |
| Nausea | 2 (13.3%) | 0 (0.0%) | 1 (7.1%) | 0 (0.0%) | 3 (6.8%) | 3 (5.7%) |
| Headache | 1 (6.7%) | 2 (13.3%) | 6 (42.9%) | 3 (33.3%) | 9 (20.5%) | 12 (22.6%) |

**Note:** Presented data are n (%) of participants experiencing each solicited symptom from vaccination through Day 8.

**SUPPLEMENTAL TABLE 6. SUMMARY OF UNSOLICITED ADVERSE EVENTS AND LABORATORY ABNORMALITIES**

| | Group A n (%) | Group B n (%) | Group C n (%) | Group D n (%) | Overall n (%) |
|---|---|---|---|---|---|
| **Description** | **Low dose** | **Medium dose** | **High dose** | **Control** | |
| Participants, n= | 15 | 15 | 14 | 10 | 54 |
| Unsolicited adverse events | | | | | |
| Participants, n= | 5 (33.3%) | 9 (60.0%) | 4 (28.6%) | 2 (20.0%) | 20 (37.0%) |
| Any events, n (%) | 8 (53.3%) | 13 (86.7%) | 5 (35.7%) | 3 (30.0%) | 29 (53.7%) |
| Any Grade 2 events, n (%) | 6 (40.0%) | 7 (46.7%) | 1 (7.1%) | 2 (20.0%) | 16 (29.6%) |
| Serious Adverse Events | | | | | |
| Participants, n= | 0 (0.0%) | 0 (0.0%) | 0 (0.0%) | 0 (0.0%) | 0 (0.0%) |
| Medically Attended Adverse Events | | | | | |
| Participants, n= | 2 (13.3%) | 1 (6.7%) | 0 (0.0%) | 2 (20.0%) | 5 (9.3%) |
| Any events, n (%) | 2 (13.3%) | 1 (6.7%) | 0 (0.0%) | 2 (20.0%) | 5 (9.3%) |
| Any Grade ≥2 events, n (%) | 2 (13.3%) | 1 (6.7%) | 0 (0.0%) | 2 (20.0%) | 5 (9.3%) |
| New-Onset Chronic Medical Conditions | | | | | |
| Participants, n= | 0 (0.0%) | 0 (0.0%) | 0 (0.0%) | 0 (0.0%) | 0 (0.0%) |
| Potential Immune-Mediated Medical Conditions | | | | | |
| Participants, n= | 0 (0.0%) | 0 (0.0%) | 0 (0.0%) | 0 (0.0%) | 0 (0.0%) |
| New-onset sensorineural hearing loss | | | | | |
| Participants, n= | 0 (0.0%) | 0 (0.0%) | 0 (0.0%) | 0 (0.0%) | 0 (0.0%) |
| Clinical laboratory adverse events | | | | | |
| Participants, n= | 2 (13.3%) | 10 (66.7%) | 5 (35.7%) | 3 (30.0%) | 20 (37.0%) |
| Any events, n (%) | 2 (13.3%) | 10 (66.7%) | 5 (35.7%) | 3 (30.0%) | 20 (37.0%) |
| Any Grade ≥2 events, n (%) | 0 (0.0%) | 5 (33.3%) | 1 (7.1%) | 0 (0.0%) | 6 (11.1%) |

**Note:** There were no unsolicited adverse events of Grade ≥3. No medically attended adverse events were determined to be related to study drug. There were no clinical laboratory adverse events of Grade ≥3. Of the 6 moderate severity clinical laboratory adverse events, 5 were determined to be related to study drug.

### SUPPLEMENTAL TABLE 7. LINE LISTING OF UNSOLICITED ADVERSE EVENTS (BLINDED)

| ID# | Description | Associated with dose # / Days since dose | Grade | Medically-attended | Relationship to study vaccine | If not related, alternative etiology | Outcome |
|---|---|---|---|---|---|---|---|
| 005 | Oropharyngeal pain | #2/ 32 | 1 | No | Not related | Concurrent illness/ condition | Recovered/ resolved |
| 007 | Dry skin | #1/ 1 | 1 | No | ***Related*** | n/a | Recovered/ resolved |
| 007 | Fibula fracture | #2/ 24 | 2 | Yes | Not related | Trauma/external factors | Not recovered/ not resolved |
| 009 | Bursitis (knee) | #1/ 7 | 2 | Yes | Not related | Concurrent illness/ condition | Not recovered/ not resolved |
| 017 | Musculoskeletal pain | #1/ 2 | 2 | No | ***Related*** | n/a | Recovered/ resolved |
| 017 | Vomiting | #1/ 2 | 2 | No | ***Related*** | n/a | Recovered/ resolved |
| 019 | Post-viral fatigue syndrome | #2/ 7 | 1 | No | Not related | Concurrent illness/ condition | Recovered/ resolved |
| 021 | URI | #2/ 3 | 2 | No | Not related | Concurrent illness/ condition | Recovered/ resolved |
| 021 | Vestibular disorder | #2/ 10 | 2 | Yes | Not related | Concurrent illness/ condition | Not recovered/ not resolved |
| 026 | Allergic rhinitis | #2/ 17 | 1 | No | Not related | Concurrent illness/ condition | Not recovered/ not resolved |
| 039 | Injection site bruising | #1/ immediate | 1 | No | Not related | Study procedure | Recovered/ resolved |
| 043 | Tachycardia | #1/ 8 | 2 | No | Not related | Other preexisting disease or condition | Recovered/ resolved |
| 044 | Intermenstrual bleeding | #2/ 0 | 1 | No | Not related | Concurrent illness/ condition | Recovered/ resolved |
| 048 | URI | #2/ 1 | 2 | No | Not related | Concurrent illness/ condition | Recovered/ resolved |
| 048 | Rhinitis | #2/ 36 | 2 | No | Not related | Concurrent Illness/ condition | Recovered/ resolved |
| 051 | URI | #1/ 2 | 1 | No | Not related | Concurrent Illness/ condition | Recovered/ resolved |
| 051 | URI | #2/ 1 | 2 | No | Not related | Concurrent Illness/ condition | Recovered/ resolved |
| 055 | URI | #1/ 19 | 2 | No | Not related | Concurrent Illness/ condition | Recovered/ resolved |
| 058 | Sinusitis | #2/ 27 | 2 | Yes | Not related | Concurrent Illness/ condition | Recovered/ resolved |
| 060 | URI | #1/ 13 | 1 | No | Not related | Concurrent Illness/ condition | Recovered/ resolved |
| 068 | Back pain | #1/ 0 | 2 | No | Not related | Concurrent Illness/ condition | Recovered/ resolved |
| 068 | URI | #2/ 10 | 1 | No | Not related | Concurrent Illness/ condition | Recovered/ resolved |
| 070 | Injection site reaction | #1/ 8 | 1 | No | ***Related*** | n/a | Not recovered/ not resolved |
| 070 | Cerumen impaction | #1/ 2 | 2 | No | Not related | Other pre-existing disease or condition | Recovered/ resolved |
| 070 | Contusion | #2/ 12 | 2 | No | Not related | Trauma or external factors | Recovered/ resolved |
| 071 | URI | #2/ 1 | 1 | No | Not related | Concurrent illness/ condition | Recovered/ resolved |
| 076 | Myalgia | #2/ 0 | 1 | No | ***Related*** | n/a | Recovered/ resolved |
| 086 | Ingrowing nail | #1/ 15 | 2 | Yes | Not related | Concurrent illness/ condition | Recovered/ resolved |
| 086 | URI | #2/ 13 | 1 | No | Not related | Concurrent illness/ condition | Recovered/ resolved |

**Note:** Study investigators remain blinded to individual treatment allocation. Grade 1=Mild, 2=Moderate. There were no Grade 3 or higher unsolicited AEs. URI=upper respiratory tract infection. There were no serious adverse events (SAEs), new-onset chronic medical conditions (NOCMCs), potentially immune-mediated medical conditions (PIMMCs), or sensorineural hearing loss. Outcome is as of Day 61; all unrecovered/unresolved adverse events will remain under monitoring until resolution.
